# Geographic Concentration of Genomic Surveillance for Highly Pathogenic Avian Influenza A(H5), South Asia, 2015–2025

**DOI:** 10.64898/2026.07.20.26358505

**Authors:** Nehal Hasnain, Selim Farhad Shihab, Md. Asadul Islam, Md. Abdur Rahman, Md. Abdul Masum

**Affiliations:** Department of Microbiology and Parasitology, Sher-e-Bangla Agricultural University (SAU), Dhaka, Bangladesh; Department of Medicine and Public Health, Sher-e-Bangla Agricultural University (SAU), Dhaka, Bangladesh; Research and Development Department, Incepta Pharmaceuticals Ltd., Dhaka, Bangladesh; Faculty of Animal Science & Veterinary Medicine, Sher-e-Bangla Agricultural University (SAU), Dhaka, Bangladesh; Department of Anatomy, Histology and Physiology, Sher-e-Bangla Agricultural University (SAU), Dhaka, Bangladesh

**Author notes:** **Corresponding author:** Md. Abdul Masum, Professor, Department of Anatomy, Histology and Physiology, Sher-e-Bangla Agricultural University (SAU), Sher-e-Bangla Nagar, Dhaka 1207, Bangladesh.

**Keywords:** . avian influenza, influenza A(H5), HPAI, genomic surveillance, molecular epidemiology, pandemic preparedness, one health, South Asia, INSDC

## Abstract

Early detection of mammalian adaptation in highly pathogenic avian influenza A(H5) depends on genomic surveillance, yet its distribution across high-burden regions is poorly characterized. We quantified open-access (GenBank/INSDC) H5 genomic coverage relative to reported outbreak burden across nine South Asian countries during 2015–2025, linking isolates to FAO EMPRES-i/WOAH events. Of 919 H5 isolates, 814 (89%) came from one country (Bangladesh); the other eight contributed 105. India, with the largest burden (322 events), yielded only 42 isolates (13 per 100); Nepal, 2 of 78; Afghanistan, none of 5. Concentration was extreme (Gini 0.83) and unchanged by adding restricted GISAID records (1,297 combined isolates; Bangladesh 89%) or by normalizing to poultry or human population. Because reported outbreaks track reporting effort, these coverage ratios are directional, not rates. This single-country dependency, deepest where burden is highest, is a regional early-warning vulnerability.

## Introduction

Highly pathogenic avian influenza (HPAI) A(H5N1) viruses of clade 2.3.4.4b have spread globally since 2021, producing the largest avian influenza panzootic on record. This has also led to an unprecedented breadth of mammalian infection, from wild carnivores and marine mammals to farmed fur animals and, in 2024, dairy cattle in North America (1–5). Each mammalian infection is an opportunity to acquire adaptations, most notably the polymerase substitutions PB2 E627K and D701N. These increase replication in mammalian cells and could move the virus toward efficient human-to-human transmission (6–8). Genomic surveillance is the principal early-warning system for these changes. Only sequence data reveal which lineages are circulating and whether mammalian-adaptive markers have emerged.

That early-warning system depends on three properties easy to assume and hard to achieve. The sequencing must be timely, the data openly accessible (for real-time analysis), and sequencing effort located where outbreaks occur. Sequences held far from the outbreak front, or in restricted archives, cannot support the rapid, distributed risk assessment a pandemic threat demand. The open-access repositories of the International Nucleotide Sequence Database Collaboration (INSDC; e.g., GenBank) are the substrate for this reuse (9–11).

These requirements matter acutely in South Asia. The region encompasses the terminal wintering grounds of the Central Asian Flyway (12), supports some of the world’s densest poultry production and live-bird-market networks, and has experienced recurrent HPAI H5 outbreaks since the mid-2000s (13–18). The resulting human–animal interface is large and persistent. However, how thoroughly H5 outbreak activity is captured in the open genomic record, and whether that characterization tracks where outbreaks occur, remain unquantified.

We therefore quantified open-access (GenBank/INSDC) H5 genomic coverage relative to reported HPAI outbreak burden across nine South Asian countries (Afghanistan, Bangladesh, Bhutan, India, Maldives, Myanmar, Nepal, Pakistan, Sri Lanka) during 2015–2025. Our aim was descriptive but consequential, to identify where, within a high-burden region, the open genomic early-warning system is strong and where it is absent, and how this should inform future sequencing capacity. Because open-access counts could understate coverage where laboratories preferentially deposit to restricted archives. We additionally tested whether the pattern persisted when GISAID EpiFlu records were added.

## Methods

### Study design

We conducted a retrospective ecological analysis of open-access influenza A(H5) genomic surveillance relative to reported HPAI outbreak burden across eight South Asian countries (Afghanistan, Bangladesh, Bhutan, India, Maldives, Nepal, Pakistan, Sri Lanka) and Myanmar for viruses collected during 2015–2025. We included Myanmar because it is contiguous with Bangladesh and India and is a Central Asian Flyway range state, making it an epidemiologically relevant extension of the South Asian study area. The country was the unit of analysis; the outcome was the ratio of genomically characterized H5 isolates to reported H5 outbreak events.

### Genomic numerator

We retrieved all influenza A records (NCBI taxonomy txid11320) for the nine countries from NCBI Nucleotide. Then confirmed each record’s origin from its “/geo_loc_name” source qualifier (NCBI exposes no indexed geographic field), restricted to collection years 2015–2025, and classified H5 from the “/serotype” qualifier (Technical Appendix). The numerator was the count of unique H5 isolates, deduplicated by “/strain” after canonicalizing strain identifiers to strip embedded segment tokens, so the eight segments of one virus collapse to a single isolate. We retained all H5 subtypes, predominantly H5N1, with H5N6, H5N8, and a few H5N2/H5N3, and report a sensitivity analysis excluding the likely low-pathogenic H5N2/H5N3 records. Clade was not resolvable from GenBank metadata and was not assigned; because the region’s H5 record spans multiple clades and predates 2021, we did not treat it as a single 2.3.4.4b sample.

### Outbreak denominator

Reported burden came from the FAO EMPRES-i Global Animal Disease Information System (19), via a single authenticated export filtered to 2015–2025 to avoid the interface’s default truncation to 15,000 global events. We counted unique H5 avian influenza outbreak events in the nine countries (deduplicated by event identifier; observation-level rows as a sensitivity), with event year from the observation date, falling back to the report date where missing. EMPRES-i ingests WOAH/WAHIS notifications (20), so the two are not independent; because the export cannot be regenerated through a public API, we archive the exact extract. To gauge the denominator’s sensitivity to its source, we separately retrieved the WOAH WAHIS epidemiological-event record for the same nine countries and period as an independent cross-check. We correlated the resulting country rank order with the EMPRES-i ranking and recomputed the coverage index under the WAHIS denominator. Because WAHIS is the coarser stream EMPRES-i ingests, this validates the ranking rather than the absolute totals (Technical Appendix).

### Coverage metric

The primary metric was unique H5 isolates per 100 reported H5 outbreak events per country. Because numerator and denominator come from two independent surveillance streams. We therefor treat this as a coverage index, not a rate. The index is unbounded and undefined where no outbreaks are reported. The denominator reflects reporting effort as much as incidence. As a result, the index interpretable directionally, not as an absolute rate. Large ratios in low-reporting countries index sparse reporting, not superior characterization. We attached exact Poisson 95% CIs to the isolate count at a fixed denominator (21). Because reported outbreaks track effort, we also summarized geographic inequality with the denominator-free Gini coefficient calculated across all nine countries (22). We normalized isolate counts by poultry-reservoir size (FAO, 2024) (23) and human population (World Bank, 2023) (24). These denominators are independent of outbreak reporting (Technical Appendix). As a pre-specified secondary metric, we recomputed coverage using only whole-genome (eight-segment) isolates, because mammalian-adaptation markers require the internal segments, not hemagglutinin alone.

### Genome completeness and host

For each isolate, we counted distinct gene segments (1 to 8) and recorded whether the PB2 segment was present. PB2 carries the mammalian-adaptation markers PB2 E627K and D701N. Its presence therefore indexes whether an isolate provides the substrate needed to screen for those markers (analytic capacity), whereas genotyping the markers themselves lay outside this metadata-based study. Host was classified from the “/host” qualifier into the following categories: domestic poultry, wild bird, genus-level *Anas* sp. of unresolved domestication status, other avian, mammal, and environment (Technical Appendix). For mammalian isolates, we additionally recorded the host species. We also noted whether the record represented a spillover or mortality detection rather than routine reservoir surveillance. Metadata completeness was the proportion of isolates with an annotated host and day-level collection-date precision.

### Deposition timeliness

To assess how quickly genomes reached the public record, we retrieved each isolate’s GenBank first-public-release date (GBSeq create-date). We did this by a targeted re-fetch of its segment accessions. We then took the isolate’s availability date as the earliest release date among its segments. The collection-to-release lag was computed the as the release date minus the collection date. The metric was restricted to the 880 of 919 isolates with a month-or-finer collection date (39 year-only dates were excluded as too coarse). We summarized the lag by country as medians with interquartile ranges (Technical Appendix).

### Robustness to restricted-database deposition

As a preregistered extension, we repeated the analysis on the union of GenBank and GISAID records. We exported all influenza A(H5) isolates (any neuraminidase subtype, any host) for the nine countries, 2015–2025, from GISAID EpiFlu. Records were deduplicating by EPI_ISL accession and cross-deduplicating against GenBank by canonical strain within country, so viruses in both databases were counted once. The denominator was unchanged (Technical Appendix). Per GISAID terms, we report only aggregate per-country counts and cite the data set (EPI_SET_260705su; doi:10.55876/gis8.260705su) with its acknowledgement table, and summarized its curated clade field.

### Reproducibility, ethics, and data availability

Analyses used Python 3.13 (pandas, Biopython, matplotlib). The study was preregistered at the Open Science Framework (https://osf.io/dcx3e; DOI 10.17605/OSF.IO/DCX3E; registered 2026-06-28); the isolate-deduplication refinement is a documented deviation from the registered rule. The concentration analysis, reservoir- and population-normalized denominators, WOAH WAHIS denominator cross-check, deposition-timeliness metric, 2015–2019/2020–2025 temporal split, and mammalian species-level characterization were exploratory additions.

All processing code, query strings, accession lists, aggregate GISAID counts, the EMPRES-i and WOAH WAHIS extracts archived at Zenodo. The FAO and World Bank denominator extracts are also archived there. A preregistered cross-region benchmark remains out of scope. The study used only publicly available sequence metadata and aggregate animal-outbreak reports. It did not involve human participants or animal experimentation and did not require institutional ethics review.

## Results

### Composition of the Open-Access Record

The open-access influenza A record for the nine countries during 2015–2025 comprised 4,324 unique isolates. Human seasonal influenza accounted for 2,017 (47%) and avian hosts for 2,130 (49%). Among avian isolates, H9 (predominantly H9N2; n=914) and H5 (n=891) co-dominated.

Across all hosts, H5 viruses totaled 919 isolates region-wide. The remainder from mammalian, environmental, and unresolved-host records. Of the H5 isolates, 806 (88%) were H5N1 and the rest H5N6 (n=40), H5N8 (n=22), H5N2 (n=16), H5N3 (n=3), and H5 of unresolved neuraminidase subtype (n=32). Excluding the 19 likely low-pathogenic H5N2/H5N3 records (all from Bangladesh and Myanmar, none from India) left the concentration and India’s deficit essentially unchanged (Bangladesh 89.2%; India 13.0 per 100; Technical Appendix section 3).

### Geographic Concentration of H5 Isolates

Of the 919 H5 isolates, 814 (89%) originated from one country, Bangladesh. The remaining eight together contributed 105 (Figure 1). This concentration was extreme by formal measure (Gini coefficient 0.83; Technical Appendix Table A5, Figure A5). It remained undiminished on the combined GenBank ∪ GISAID record (Gini coefficient 0.83). Bangladesh’s deposition was sustained rather than historical, with 2023–2025 among its highest-volume years.

**Figure 1.**
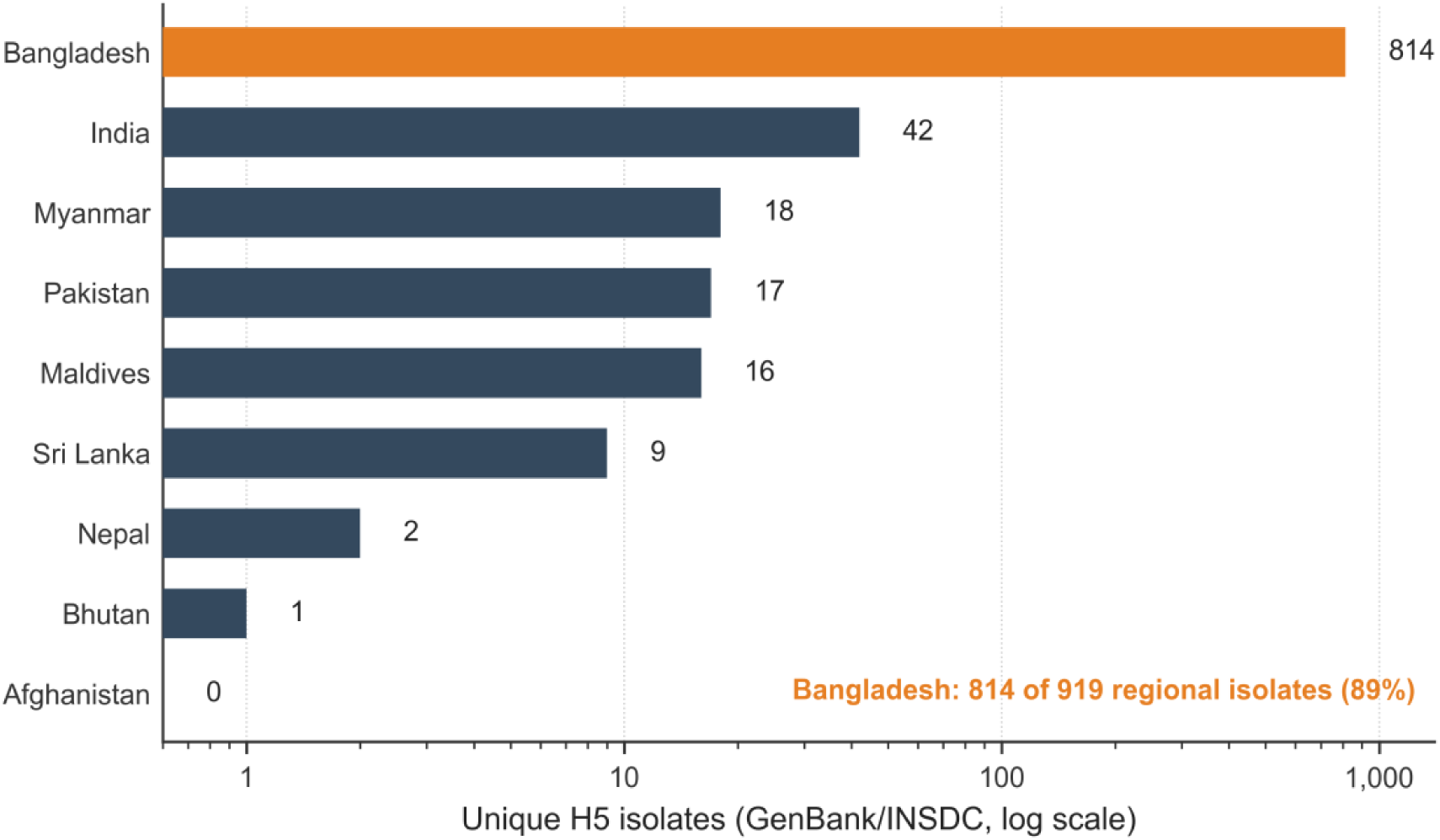
Open-access influenza A(H5) isolates by country, South Asia, 2015–2025. **Note:** Bars show the number of unique H5 isolates (GenBank/INSDC, deduplicated to the isolate level) contributed by each of the nine countries, plotted on a logarithmic x-axis so that all nine countries remain legible despite Bangladesh’s dominance; Afghanistan (0 isolates) is shown at the axis floor. Bangladesh (highlighted in orange) contributed 814 of the 919 regional isolates (89%); the other eight countries together contributed 105.

### Coverage Relative to Reported Outbreaks

Genomic coverage carries little relation to where outbreaks were reported (Table 1; Figure 2). India reported the largest H5 outbreak burden (322 events), yet yielded only 42 isolates. This is about one sequence for every eight reported outbreaks. This gives a coverage index of 13.0 per 100 (95% CI 9.4–17.6). This is roughly an order of magnitude below the one-per-outbreak parity that other reporting countries exceed.

**Figure 2.**
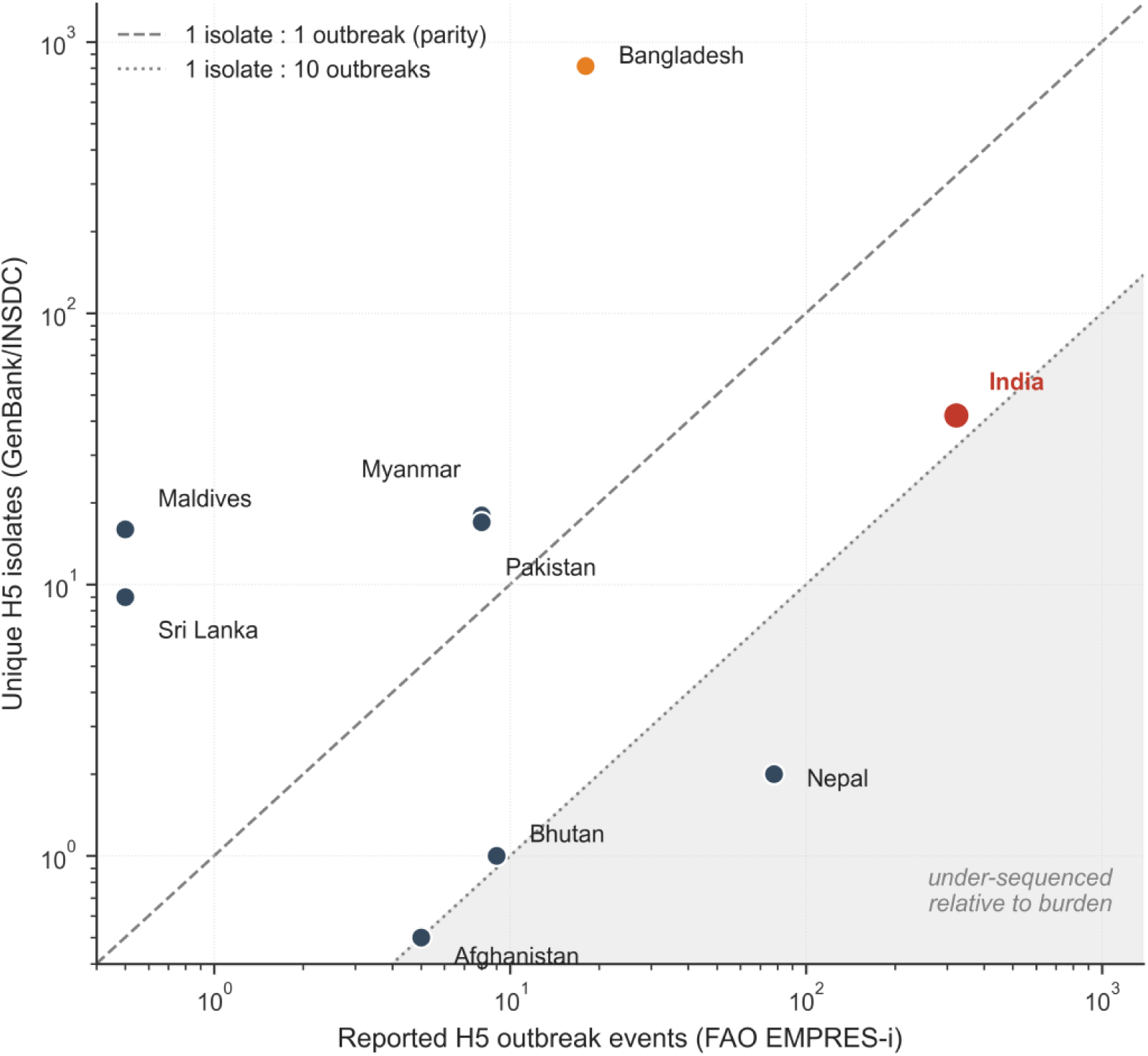
Genomic coverage versus reported outbreak burden, South Asia, 2015–2025. Each point is one country, plotted by reported H5 outbreak events (FAO EMPRES-i, x-axis) against unique H5 isolates (GenBank/INSDC, y-axis) on logarithmic axes. The dashed line marks 1 isolate per outbreak (parity) and the dotted line 1 isolate per 10 outbreaks; the shaded region below indicates under-sequencing relative to reported burden. India (red), the highest-burden country, lies deep in the under-sequenced region, whereas Bangladesh (orange) lies far above parity. Countries reporting no outbreaks (Maldives, Sri Lanka) are plotted at a floor of 0.5 for display on logarithmic axes.

**Table 1.** Open-access H5 genomic coverage and metadata completeness, by country, South Asia, 2015–2025. (unique open-access H5 isolates, canonical dedup; ordered by reported outbreak burden)

| Country | Isolates | Outbreaks | Iso/100 | 95% CI | Poultry | Wild | *Anas sp.* | Mammal | Host known % | Day-precise % |
| --- | --- | --- | --- | --- | --- | --- | --- | --- | --- | --- |
| India | 42 | 322 | 13.0 | 9.4–17.6 | 17 | 6 | 9 | 9 | 100 | 81.0 |
| Nepal | 2 | 78 | 2.6 | 0.3–9.3 | 2 | 0 | 0 | 0 | 100 | 100 |
| Bangladesh | 814 | 18 | 4522.2 | 4217–4844 | 222 | 139 | 436 | 1 | 99.5 | 99.9 |
| Bhutan | 1 | 9 | 11.1 | 0.3–61.9 | 1 | 0 | 0 | 0 | 100 | 0 |
| Myanmar | 18 | 8 | 225.0 | 133–356 | 4 | 0 | 0 | 0 | 100 | 22.2 |
| Pakistan | 17 | 8 | 212.5 | 124–340 | 16 | 0 | 0 | 0 | 94.1 | 94.1 |
| Afghanistan | 0 | 5 | 0 | 0–73.8 | 0 | 0 | 0 | 0 | — | — |
| Maldives | 16 | 0 | — | — | 0 | 0 | 0 | 0 | 100 | 0 |

| Country | Isolates | Outbreaks | Iso/100 | 95% CI | Poultry | Wild | Anas sp. | Mammal | Host known % | Day-precise % |
| --- | --- | --- | --- | --- | --- | --- | --- | --- | --- | --- |
| Sri Lanka | 9 | 0 | — | — | 0 | 0 | 0 | 0 | 100 | 0 |

**Note:** Iso/100 is a coverage index (unique H5 isolates per 100 reported outbreak events), with an exact Poisson (gamma-method) 95% CI on the isolate count; the interval reflects the discreteness of the genomic count, not reporting uncertainty in the denominator, and is undefined where no outbreaks were reported. Avian-unspecified, environment, and unknown host classes omitted from the per-country columns for space (region-wide 4.2% / 0.3% / 1.6%); full breakdown in outputs/host\_stratification\_\*.csv. Day-precise % is unstable where n is small (Bhutan, Maldives, Sri Lanka have 1–16 isolates).

Coverage was lower still in Nepal (2 per 78; 2.6) and Afghanistan (none for 5 reported events), and modest in Bhutan (1 per 9). Countries reporting few outbreaks showed high ratios. Examples include Bangladesh (814 per 18 events), Myanmar (225 per 100), and Pakistan (212 per 100). These high ratios reflect sparse reporting, not superior characterization. Maldives and Sri Lanka contributed isolates (16 and 9) despite reporting no outbreaks.

Even the upper bound of India’s interval (17.6 per 100) fell below the lower bounds of the high-coverage countries (Bangladesh, 4,217; Myanmar, 133; Pakistan, 124). Thus, its deficit is not an artifact of a small genomic count. although the index’s magnitude is denominator-dependent (13 per 100 under EMPRES-i vs 58 under WAHIS; Table A1), India stayed below parity under either denominator.

### Combined GenBank and GISAID Coverage

Integrating the 1,149 regional GISAID EpiFlu isolates with GenBank yielded 1,297 unique isolates after cross-deduplication. Of those, 717 of Bangladesh’s were present in both databases. Concentration was unchanged: Bangladesh accounted for 1,158 of 1,297 (89%), the other eight countries for 139 (Table 2).

**Table 2.** Combined open-access (GenBank) and GISAID H5 coverage, South Asia, 2015–2025.

| Country | GenBank | GISAID | In both | Combined | Outbreaks | Combined iso/100 (95% CI) |
| --- | --- | --- | --- | --- | --- | --- |
| India | 42 | 41 | 37 | 46 | 322 | 14.3 (10.5–19.1) |
| Nepal | 2 | 5 | 2 | 5 | 78 | 6.4 (2.1–15.0) |
| Bangladesh | 814 | 1,061 | 717 | 1,158 | 18 | 6433 (6068–6815) |
| Bhutan | 1 | 1 | 1 | 1 | 9 | 11.1 (0.3–61.9) |
| Myanmar | 18 | 11 | 4 | 25 | 8 | 312 (202–461) |
| Pakistan | 17 | 14 | 10 | 21 | 8 | 262 (162–401) |
| Afghanistan | 0 | 0 | 0 | 0 | 5 | 0 (0–73.8) |
| Maldives | 16 | 11 | 0 | 27 | 0 | — |
| Sri Lanka | 9 | 5 | 0 | 14 | 0 | — |
| Region | 919 | 1,149 | 771 | 1,297 | 448 | — |

**Note:** Unique isolates after cross-deduplication; ordered by reported outbreak burden. GISAID records shared under EPI\_SET\_260705su (doi:10.55876/gis8.260705su); only aggregate counts are reported, per GISAID data-sharing
terms. “In both” = isolates present in GenBank and GISAID and counted once in the combined column; combined = GenBank ∪ GISAID unique isolates.

India’s deficit was not an artifact of open-access-only counting. GISAID held only 41 India isolates, just 4 additional to GenBank. This raised India’s count from 42 to 46, and its index 13.0 to 14.3 per 100. Combined coverage stayed below 7 per 100 in Nepal (5 of 78; 6.4) and zero in Afghanistan. Maldives and Sri Lanka again contributed isolates (27 and 14) despite no reported outbreaks.

### Coverage by Poultry and Population

We also re-expressed coverage per unit of poultry reservoir and per capita (Technical Appendix Table A6, Figure A6). Bangladesh again dominated, with 2,055 isolates per billion poultry and 47 per 10 million people. This confirms genuine sequencing intensity rather than a low-reporting artifact.

By contrast, India (47 per billion poultry) and Pakistan (8) lay near the floor. This was despite holding the region’s two largest flocks (0.9 and 2.1 billion birds). Pakistan’s apparently high outbreak-based index (212 per 100) reflected sparse reporting rather than dense sequencing. Relative to its reservoir, it was the region’s most under- sampled country.

### H5 Clade Composition

Among the 1,149 regional GISAID isolates, 1,069 carried a curated hemagglutinin clade and 80 were unassigned. The older enzootic 2.3.2.1a predominated (639), ahead of the panzootic 2.3.4.4b (329). The remainder were distributed across 2.3.4.4h (42), 2.3.2.1c (25), and several minor clades.

Because these isolates are overwhelmingly Bangladeshi, this composition principally describes Bangladesh’s long-established H5N1 record. However, it confirms that the open H5 record cannot be treated as a single 2.3.4.4b sample. It also leaves India and other thinly sequenced countries largely clade uncharacterized.

### Whole-Genome Availability

Only 526 of 919 isolates (57%) were whole genomes. A further 213 (23%) comprised a single segment. PB2, required to screen for mammalian-adaptation markers, was present in 71% (Figure 3).

**Figure 3.**
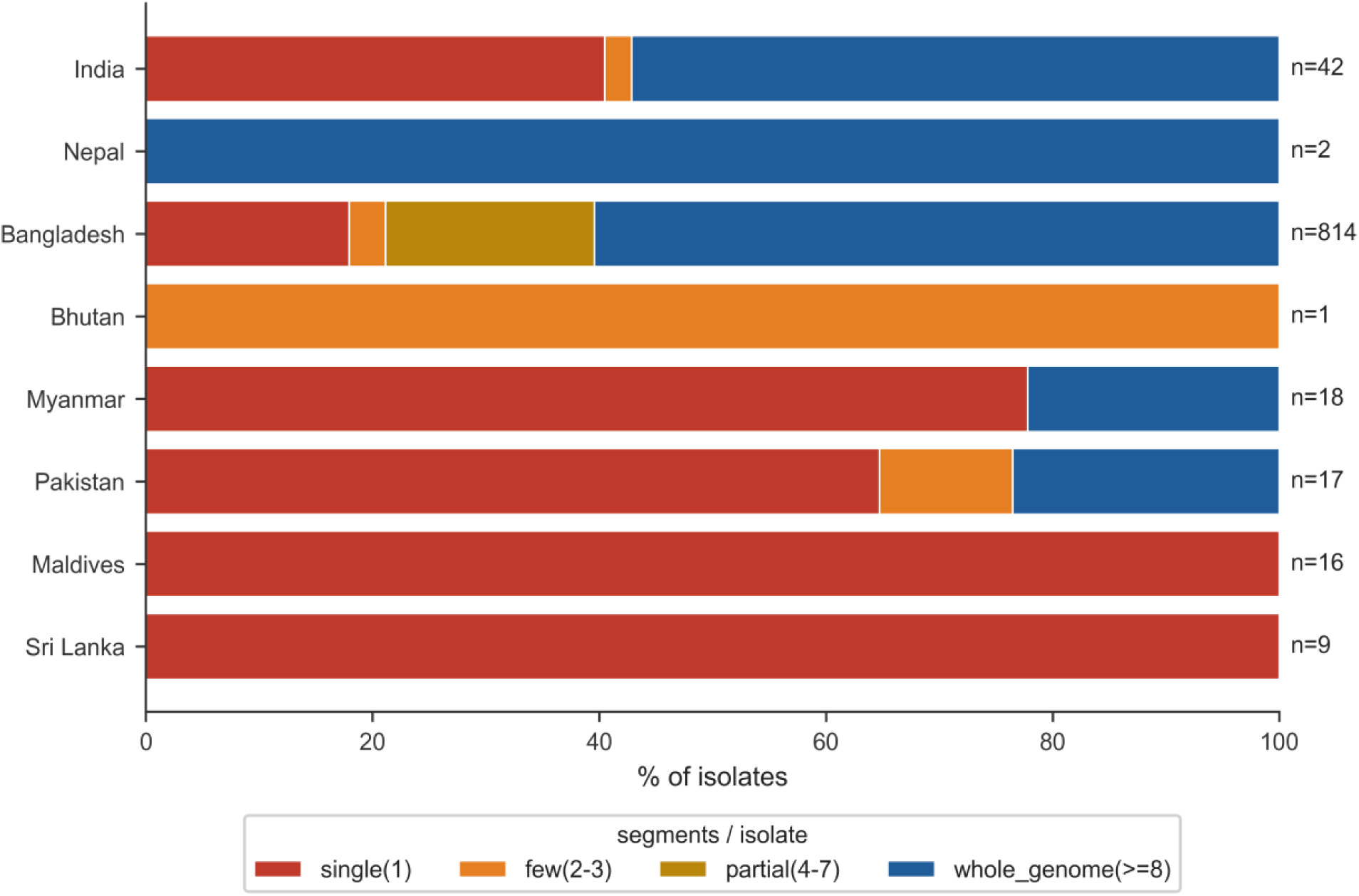
Genome completeness of open-access H5 isolates by country, South Asia, 2015–2025 (n=919). Horizontal bars show, for each country, the percentage of isolates by number of gene segments deposited: single segment (1), few (2–3), partial (4–7), and whole genome (≥8); per-country isolate counts (n) are shown at right. Whole-genome fractions were 60% in Bangladesh and 57% in India but 0% in Bhutan, Maldives, and Sri Lanka, whose deposits were limited to the hemagglutinin and neuraminidase genes.

Completeness was geographically structured. Whole-genome fractions of 60% in Bangladesh and 57% in India, but 22–24% in Myanmar and Pakistan. In Bhutan, Maldives, and Sri Lanka, the whole-genome fraction was 0%. Deposits from these three countries were limited to hemagglutinin and neuraminidase genes and contained no PB2. Restricting the coverage metric to whole genomes widened the gap further. India’s index fell from 13.0 to 7.5 whole genomes per 100 reported outbreaks.

### Sequence-Release Timeliness

The open record was also slow. Across the 880 of 919 isolates with a month-or-finer collection date, the median collection-to-release lag was 392 days (∼13 months; IQR 238–762). This exceeded a year even in the two countries that sequence at scale (Bangladesh, 377 days; India, 494; Technical Appendix Table A4).

Here, “release” means an isolate’s first public availability in GenBank (the earliest release date among its segments), not its sequencing or manuscript-submission date. A genome released more than a year after the sampled event is a retrospective archive, not a real-time signal.

### Metadata Completeness and Host Distribution

Host was annotated for >99% of isolates, and dates were day-level for 95%, none missing (Table 1). The deficit was one of volume, not curation. By host, the record was dominated by domestic poultry (28.5%) and genus-level *Anas sp*. waterfowl (48.4%, almost entirely Bangladesh). Wild birds accounted for 15.8% and mammals 1.1%.

### Mammalian H5 Detections

Ten H5 isolates derived from mammals (four tigers, three domestic cats, two leopards, and one human), all collected during 2021–2025. Nine, including the single human isolate and every big-cat detection, came from India. These constituted 9 of India’s 42 isolates (21%). About one-fifth of India’s open-access H5 record reflected spillover or mortality events rather than routine reservoir surveillance.

## Discussion

Across nine South Asian countries and eleven years, open-access genomic surveillance for HPAI A(H5) was extremely concentrated in a single country. To the extent that reported outbreaks approximate true burden, it was also decoupled from that burden.

Nearly nine in ten regional H5 isolates (814 of 919; 89%) came from Bangladesh. This concentration is extreme by formal measure (Gini coefficient 0.83). India, despite the region’s largest reported outbreak burden (322 events), contributed only 42 (13 per 100). Other reporting countries contributed almost nothing (Nepal, 2 of 78; Afghanistan, 0 of 5; Bhutan, 1 of 9). This pattern was not an artifact of open-access counting, as adding the restricted GISAID EpiFlu record left the concentration essentially unchanged (Bangladesh, 89% of 1,297 combined isolates). India’s deficit also remain intact (46 isolates; 14 per 100).

That dependency is current, not historical. Bangladesh’s deposition continued through 2023–2025 among its highest annual volumes and increasingly as whole genomes. The concentration therefore describes the system as it operates today. An interruption to that one national effort, whether in funding, laboratory capacity, or data-sharing policy, would darken the open genomic picture region-wide. A surveillance architecture for a pathogen of pandemic potential should not contain such a single point of failure. It is also more precarious than a raw count implies. The open regional record derives substantially from a few externally supported research collaborations centered on Bangladesh (15,16,18), rather than routine national deposition. This is not an alternative explanation to argue away, but the vulnerability itself. Prior work documented genomic-surveillance inequity between income strata: only 42% of low- and middle-income countries sequenced >0.5% of cases during COVID-19, versus 78% of high-income countries (25,26). The pattern here differs in kind. It showes an extreme intra-regional concentration in which one lower-middle-income country supplies 89% of the record, while the region’s highest-burden country stays nearly unsequenced. To our knowledge, this dependency has not been previously quantified for HPAI H5.

India appears to be an important gap in the accessible record, and its pattern is informative in a second respect. Nine of India’s 42 open-access isolates (21%) derive from mammals. These include the region’s only sequenced human H5 isolate and every big-cat detection, all from the panzootic era (2021–2025). The limited number of isolates available through the open-access record is weighted toward reported spillovers and die-offs, while the extent to which the poultry reservoir that drives transmission has been sampled and sequenced is difficult to assess from these data (14,27). The presence of only four isolates from India in the restricted GISAID dataset may reflect limited submission to that repository, deposition in other or national databases, gaps in sequencing, or some combination of these factors. The present study cannot distinguish among these possibilities. The observed pattern may be consistent with an animal-health system in which outbreak response emphasizes rapid control measures, with routine genomic characterization varying across outbreaks and settings (28). These findings suggest the value of strengthening routine genomic surveillance and data sharing. Early warning may be less complete precisely where the stakes, including detection of mammalian adaptation are high (29). India’s coverage ratio has in fact risen over the study period, so the gap is not widening. Nevertheless, India remains under-covered in the accessible dataset. These interpretations are inferential. The ecological, record-based design can identify the coverage gap but not establish its cause, because no farm-level, laboratory-capacity, or funding data were within the study’s scope.

The gap is also deeper than isolate counts imply. Only 57% of regional H5 isolates were whole genomes. PB2, which carries the mammalian-adaptation markers surveillance exists to detect, was present in just 71%. This deficit is geographically structured. Isolates outside Bangladesh and India were largely limited to hemagglutinin and neuraminidase. Those from Bhutan, Maldives, and Sri Lanka contained no PB2 at all. Restricting the coverage metric to whole genomes widened the gap further, lowering India from 13.0 to 7.5 analyzable genomes per 100 reported outbreaks. Crucially, we quantified whether the substrate for marker surveillance exists, not the markers themselves. Screening the PB2-bearing isolates for E627K and D701N is a sequence-level analysis beyond this metadata-based assessment and remains the necessary next step wherever PB2 is available.

Encouragingly, the deficit is one of volume and completeness, not data quality. Where isolates existed, they were well annotated (host >99%, day-level dates 95%, none missing). The corrective is therefore not better metadata practice but more sequencing, particularly whole-genome sequencing, and timely open deposition, distributed toward the high-burden, low-coverage settings identified here. India should be prioritized first, followed by Pakistan and Nepal (both thin relative to their poultry reservoirs), and Afghanistan, which has contributed none. Priority should go to routine reservoir (poultry and waterfowl) sampling and whole-genome deposition (26), so the open record reflects ongoing circulation rather than only its most visible consequences. Existing multilateral mechanisms, including the FAO–WOAH OFFLU network (30) and the Quadripartite One Health partnership (31), already coordinate reference-laboratory capacity and could prioritize this effort. Such efforts must, however, engage with why laboratories sometimes prefer restricted archives. Open deposition can be perceived to forgo the benefit-sharing protections that instruments such as the Pandemic Influenza Preparedness Framework and the WHO Pandemic Agreement are meant to guarantee (32–35). Sustainable coverage will depend as much on equitable access-and-benefit-sharing as on sequencing capacity itself.

Several limitations qualify these findings. Most important, the denominator reflects reporting effort, not true outbreak burden. The high coverage ratios of Bangladesh, Pakistan, and Myanmar partly reflect under-reporting of outbreaks to international systems (36–38) rather than superior characterization. This bias is directional and runs counter to the finding it might appear to threaten. Under-ascertainment shrinks the denominator and inflates the ratio, so India’s 13 isolates per 100 reported outbreaks is an upper bound on its true coverage. Fuller ascertainment would only enlarge the deficit. The high ratios should not be over-read for the same reason. Bangladesh’s 18 recorded events over eleven years is implausibly low for a country with endemic H5N1 and intensive duck and live-bird-market surveillance. An independent cross-check against WOAH WAHIS notifications reproduced the rank order almost exactly (Spearman ρ = 0.99), with India again recording the most H5 events (73; 64% of the total). Because WAHIS is the coarser stream EMPRES-i ingests, the check validates the ranking rather than the totals. Both outbreak denominators nonetheless descend from the same WOAH/WAHIS notification pipeline, so neither is an independent measure of true incidence. It is the reservoir- and population-normalized denominators, enumerated outside animal-health reporting entirely, that establish the gap is not a reporting artifact, nor is sampling error the explanation. India’s exact Poisson interval (9.4–17.6 per 100) excludes the high-coverage countries entirely. The narrow intervals around the high ratios (e.g., Bangladesh, 4,217–4,844) locate their true uncertainty in the reported-outbreak denominator, not the genomic count.

Most reassuringly, the coverage gap did not depend on the outbreak denominator. Normalizing genomic output instead by poultry-reservoir size or human population, both enumerated independently of outbreak reporting, left Bangladesh’s dominance and the thin records of India and Pakistan intact. It also exposed the high outbreak-based ratios of Pakistan and Myanmar as reporting artifacts (Technical Appendix Table A6). The FAO poultry stocks anchoring this comparison are, if anything, a conservative proxy for India, whose large backyard flock is difficult to enumerate fully. Any larger true reservoir would only deepen its deficit. The concentration result is immune to the caveat altogether, being a property of the genomic record alone.

Second, our primary numerator is open-access only, undercounting coverage wherever laboratories prefer the restricted GISAID EpiFlu database (39,40). Integrating the region’s GISAID records left the concentration and India’s deficit intact (Table 2), so the pattern is not an artifact of database choice. This inference is strongest for India, whose count rose by only four isolates. For Nepal, Pakistan and Afghanistan the restricted record is similarly sparse but too small to exclude a database-choice contribution with equal confidence. We make no cross-region comparison, having not assembled comparably GISAID-complete numerators elsewhere (25).

Third, domestication status was indeterminate for the 48% of isolates with genus-level *Anas* sp. labels (almost all Bangladeshi duck surveillance), reported separately. These almost certainly represent domestic-duck reservoir surveillance in Bangladesh’s wetlands and live-bird markets (16,18) rather than wild-bird sampling. However, the genus-level annotation precludes a definitive wild-versus-domestic assignment. Because the category is nearly confined to one country, it does not extend reservoir characterization to the under-covered countries.

Fourth, GenBank serotype does not encode pathotype; the ∼2% of likely low-pathogenic H5N2/H5N3 isolates were retained and disclosed. Excluding these 19 isolates (all in Bangladesh and Myanmar, none in India) left the concentration and India’s deficit unchanged (Bangladesh 89.2% of the record; India 13.0 per 100; Technical Appendix section 3). We did not clade-type the open-access records, though GISAID annotations confirmed a multi-clade composition dominated by the enzootic 2.3.2.1a lineage. Finally, 2024–2025 counts are floors owing to deposition lag. Deposition continuing into early 2026 (including seven further Indian H5 isolates) falls outside the preregistered window and was excluded.

Even with these caveats, the central message is robust and actionable. The open genomic early-warning system for HPAI H5 in South Asia depends on one country’s effort. It is thinnest in the highest-burden country and shallowest in genome completeness where it most needs depth. It also reaches the public record too slowly, at a median 13-month delay after collection, to support real-time response. A region defined by a major migratory flyway, dense poultry production, and an extensive human–animal interface cannot rely on the voluntary deposition of a single national program. Real-time response requires data that can be freely redistributed as outbreaks unfold, increasingly central to frameworks for pathogen access and benefit-sharing (32–35). Quantifying where the open record is absent is the prerequisite to closing those gaps before, not after, the next mammalian-adaptation event.

## Acknowledgments

We gratefully acknowledge the authors and the originating and submitting laboratories that generated and deposited the influenza A(H5) sequences used in this study to GenBank/INSDC and to the GISAID EpiFlu database; the GISAID data set analyzed here is available under identifier EPI_SET_260705su (doi:10.55876/gis8.260705su), and the corresponding acknowledgment table listing all originating and submitting laboratories accompanies the archived data. We also acknowledge the Food and Agriculture Organization of the United Nations (EMPRES-i) and the World Organisation for Animal Health (WOAH/WAHIS) for the animal-disease outbreak data used to construct and cross-check the outbreak denominator.

## Data availability

All processing code, query strings, the GenBank accession list, aggregate GISAID counts (EPI_SET_260705su; doi:10.55876/gis8.260705su) with the acknowledgement table, the FAO EMPRES-i export (filtered to the nine study countries), and the WOAH WAHIS event extract are archived at Zenodo (doi: 10.5281/zenodo.21456762)

## Author contributions

N.H. and M.A.M designed the study. N.H. and S.F.S. performed the analyses and drafted the manuscript. M.A.M, M.A.R. and M.A.I. provided critical input on the analysis and the drafted manuscript. All authors read and approved the final manuscript.

## Use of artificial intelligence

The authors used a generative AI assistant to refine the language of the manuscript. All AI-assisted text was critically reviewed by the authors, who take full responsibility for the manuscript’s content and integrity. The tool was not used as a source of scientific data or interpretation and is not listed as an author, in accordance with ICMJE guideline.

## References

1. Peacock TP, Moncla L, Dudas G, VanInsberghe D, Sukhova K, Lloyd-Smith JO, et al. The global H5N1 influenza panzootic in mammals. Nature. 2025;637:304–13. doi:10.1038/s41586-024-08054-z.

2. Agüero M, Monne I, Sánchez A, Zecchin B, Fusaro A, Ruano MJ, et al. Highly pathogenic avian influenza A(H5N1) virus infection in farmed minks, Spain, October 2022. Euro Surveill. 2023;28:2300001. doi:10.2807/1560-7917.ES.2023.28.3.2300001.

3. Leguia M, Garcia-Glaessner A, Muñoz-Saavedra B, Juarez D, Barrera P, Calvo-Mac C, et al. Highly pathogenic avian influenza A (H5N1) in marine mammals and seabirds in Peru. Nat Commun. 2023;14:5489. doi:10.1038/s41467-023-41182-0.

4. Caserta LC, Frye EA, Butt SL, Laverack M, Nooruzzaman M, Covaleda LM, et al. Spillover of highly pathogenic avian influenza H5N1 virus to dairy cattle. Nature. 2024;634:669–76. doi:10.1038/s41586-024-07849-4.

5. Burrough ER, Magstadt DR, Petersen B, Timmermans SJ, Gauger PC, Zhang J, et al. Highly Pathogenic Avian Influenza A(H5N1) Clade 2.3.4.4b Virus Infection in Domestic Dairy Cattle and Cats, United States, 2024. Emerg Infect Dis. 2024;30:1335–43. doi:10.3201/eid3007.240508.

6. Li C, Lyu X, Li X, Jia Z, Chai H. Current Status of Clade 2.3.4.4b H5N1 Highly Pathogenic Avian Influenza Virus Transmission in Mammals. China CDC Wkly. 2026;8:222–8. doi:10.46234/ccdcw2026.027.

7. Mohammad I, Hajelbashir MI, El-Bidawy MH, Abuderman A, Satea M, Arafah AMR, et al. Deciphering HPAI Influenza A Virus (H5N1): Molecular Basis of Pathogenicity, Zoonotic Potential, and Advances in Vaccination Strategies. Viruses. 2026;18:410. doi:10.3390/v18040410.

8. Wang W, Xing J, Jiang H, Lu F, Huang H, Zhang Y, et al. Human infections with avian influenza A(H5) viruses with potential pandemic risk: 1997-2025. Natl Sci Rev. 2026;13:nwaf471. doi:10.1093/nsr/nwaf471.

9. Timme RE, Karsch-Mizrachi I, Waheed Z, Arita M, MacCannell D, Maguire F, et al. Putting everything in its place: using the INSDC compliant Pathogen Data Object Model to better structure genomic data submitted for public health applications. Microb Genom. 2023;9:mgen001145. doi:10.1099/mgen.0.001145.

10. Hadfield J, Megill C, Bell SM, Huddleston J, Potter B, Callender C, et al. Nextstrain: real-time tracking of pathogen evolution. Bioinformatics. 2018;34:4121–3. doi:10.1093/bioinformatics/bty407.

11. Black A, MacCannell DR, Sibley TR, Bedford T. Ten recommendations for supporting open pathogen genomic analysis in public health. Nat Med. 2020;26:832–41. doi:10.1038/s41591-020-0935-z.

12. Global Consortium for H5N8 and Related Influenza Viruses. Role for migratory wild birds in the global spread of avian influenza H5N8. Science. 2016;354:213–7. doi:10.1126/science.aaf8852.

13. Sims LD, Domenech J, Benigno C, Kahn S, Kamata A, Lubroth J, et al. Origin and evolution of highly pathogenic H5N1 avian influenza in Asia. Vet Rec. 2005;157:159–64. doi:10.1136/vr.157.6.159.

14. Dhingra MS, Dissanayake R, Negi AB, Oberoi M, Castellan D, Thrusfield M, et al. Spatio-temporal epidemiology of highly pathogenic avian influenza (subtype H5N1) in poultry in eastern India. Spat Spatiotemporal Epidemiol. 2014;11:45–57. doi:10.1016/j.sste.2014.06.003.

15. Barman S, Turner JCM, Kamrul Hasan M, Akhtar S, Jeevan T, Franks J, et al. Emergence of a new genotype of clade 2.3.4.4b H5N1 highly pathogenic avian influenza A viruses in Bangladesh. Emerg Microbes Infect. 2023;12:e2252510. doi:10.1080/22221751.2023.2252510.

16. Turner JCM, Barman S, Feeroz MM, Hasan MK, Akhtar S, Walker D, et al. Distinct but connected avian influenza virus activities in wetlands and live poultry markets in Bangladesh, 2018-2019. Transbound Emerg Dis. 2022;69:e605–20. doi:10.1111/tbed.14450.

17. Khan SU, Berman L, Haider N, Gerloff N, Rahman MZ, Shu B, et al. Investigating a crow die-off in January-February 2011 during the introduction of a new clade of highly pathogenic avian influenza virus H5N1 into Bangladesh. Arch Virol. 2014;159:509–18. doi:10.1007/s00705-013-1842-0.

18. Barman S, Marinova-Petkova A, Hasan MK, Akhtar S, El-Shesheny R, Turner JC, et al. Role of domestic ducks in the emergence of a new genotype of highly pathogenic H5N1 avian influenza A viruses in Bangladesh. Emerg Microbes Infect. 2017;6:e72. doi:10.1038/emi.2017.60.

19. Food and Agriculture Organization of the United Nations. EMPRES-i: Global Animal Disease Information System [database]. Rome: The Organization; 2024 [cited 2026 Jul 5]. https://empres-i.apps.fao.org.

20. Caceres P, Awada L, Weber-Vintzel L, Morales R, Meske M, Tizzani P. The World Animal Health Information System as a tool to support decision-making and research in animal health. Rev Sci Tech. 2023;42:242–51. doi:10.20506/rst.42.3367.

21. Garwood F. Fiducial limits for the Poisson distribution. Biometrika. 1936;28:437–42. doi:10.1093/biomet/28.3-4.437.

22. Gini C. Measurement of inequality of incomes. Econ J. 1921;31:124–5. doi:10.2307/2223319.

23. Food and Agriculture Organization of the United Nations. Live animals: poultry stocks, 2024 [dataset]. FAOSTAT. Rome: The Organization; 2024. Distributed by Our World in Data [cited 2026 Jul 5]. https://ourworldindata.org/grapher/poultry-livestock-count.

24. World Bank. Population, total (SP.POP.TOTL) [dataset]. World Development Indicators. Washington (DC): The World Bank; 2023 [cited 2026 Jul 5]. https://data.worldbank.org/indicator/SP.POP.TOTL.

25. Brito AF, Semenova E, Dudas G, Hassler GW, Kalinich CC, Kraemer MUG, et al. Global disparities in SARS-CoV-2 genomic surveillance. Nat Commun. 2022;13:7003. doi:10.1038/s41467-022-33713-y.

26. Ohlsen EC, Hawksworth AW, Wong K, Guagliardo SAJ, Fuller JA, Sloan ML, et al. Determining Gaps in Publicly Shared SARS-CoV-2 Genomic Surveillance Data by Analysis of Global Submissions. Emerg Infect Dis. 2022;28:S85–92. doi:10.3201/eid2813.220780.

27. Kumar M, Nagarajan S, Murugkar HV, Saikia B, Singh B, Mishra A, et al. Emergence of novel reassortant H6N2 avian influenza viruses in ducks in India. Infect Genet Evol. 2018;61:20–3. doi:10.1016/j.meegid.2018.03.005.

28. Department of Animal Husbandry and Dairying, Ministry of Fisheries, Animal Husbandry and Dairying, Government of India. Action plan for prevention, control and containment of avian influenza (revised 2021). New Delhi: The Department; 2021 [cited 2026 Jul 5]. https://dahd.gov.in.

29. Sood R, Kumar N, Gokhe SS, Pateriya AK, Bhat S, Bhatia S, et al. Identification and molecular characterization of H9N2 viruses carrying multiple mammalian adaptation markers in resident birds in central-western wetlands in India. Infect Genet Evol. 2021;94:105005. doi:10.1016/j.meegid.2021.105005.

30. Edwards S. OFFLU network on avian influenza. Emerg Infect Dis. 2006;12:1287–8. doi:10.3201/eid1208.060380.

31. Food and Agriculture Organization of the United Nations, United Nations Environment Programme, World Health Organization, World Organisation for Animal Health. One Health joint plan of action (2022–2026): working together for the health of humans, animals, plants and the environment. Rome: FAO; 2022. doi:10.4060/cc2289en.

32. Eurosurveillance editorial team. Agreement on a pandemic influenza preparedness framework for the sharing of viruses and benefit sharing. Euro Surveill. 2011;16:19851.

33. Rourke MF. Access by Design, Benefits if Convenient: A Closer Look at the Pandemic Influenza Preparedness Framework’s Standard Material Transfer Agreements. Milbank Q. 2019;97:91–112. doi:10.1111/1468-0009.12364.

34. Radeino Ambe J, Bhan A, Silaigwana B, Salas SP, Edwards S, Ethics Working Group of the Coalition for Equitable Research in Low Resource Settings (CERCLE). Pathogen access and benefit sharing in a pandemic: working towards fair exchange? Lancet Microbe. 2026;7:101376. doi:10.1016/j.lanmic.2026.101376.

35. Chen L. Pathogen Access and Benefit-Sharing: Can the WHO Pandemic Agreement Bridge the Equity Divide? China CDC Wkly. 2026;8:55–7. doi:10.46234/ccdcw2026.009.

36. Delabouglise A, Antoine-Moussiaux N, Phan TD, Dao DC, Nguyen TT, Truong BD, et al. The Perceived Value of Passive Animal Health Surveillance: The Case of Highly Pathogenic Avian Influenza in Vietnam. Zoonoses Public Health. 2016;63:112–28. doi:10.1111/zph.12212.

37. Delabouglise A, Thanh NTL, Xuyen HTA, Nguyen-Van-Yen B, Tuyet PN, Lam HM, et al. Poultry farmer response to disease outbreaks in smallholder farming systems in southern Vietnam. Elife. 2020;9:e59212. doi:10.7554/eLife.59212.

38. Govindaraj G, Sridevi R, Nandakumar SN, Vineet R, Rajeev P, Binu MK, et al. Economic impacts of avian influenza outbreaks in Kerala, India. Transbound Emerg Dis. 2018;65:e361–72. doi:10.1111/tbed.12766.

39. Shu Y, McCauley J. GISAID: Global initiative on sharing all influenza data - from vision to reality. Euro Surveill. 2017;22:30494. doi:10.2807/1560-7917.ES.2017.22.13.30494.

40. Elbe S, Buckland-Merrett G. Data, disease and diplomacy: GISAID’s innovative contribution to global health. Glob Chall. 2017;1:33–46. doi:10.1002/gch2.1018

