## Supplementary Appendix for "Geographic Concentration of Genomic Surveillance for Highly Pathogenic Avian Influenza A(H5), South Asia, 2015–2025"

### Technical Appendix

This appendix provides procedural detail supporting the Methods. All processing code, query strings, the GenBank accession list, aggregate GISAID counts (`EPI_SET_260705su; doi:10.55876/gis8.260705su`), the FAO EMPRES-i export, and the WOAH WAHIS event extract (retrieved 2026-07-05) are archived at Zenodo.

#### 1. Geographic attribution of GenBank records

NCBI Nucleotide exposes no indexed country field: a country term entered as a search qualifier silently falls back to a free-text search that also matches author affiliations and cited references, so a naive query over-attributes records to well-published countries. We therefore retrieved all influenza A virus records (NCBI `taxonomy txid11320`) per country and confirmed the origin of every record from its “`/geo_loc_name`” source qualifier (or the legacy “`/country`” qualifier), retaining only records whose parsed qualifier matched the target country. Records without a resolvable country qualifier were excluded. Records were then restricted to a collection year of 2015–2025 parsed from the “`/collection_date`” qualifier.

#### 2. Isolate deduplication and strain canonicalization

Influenza A has a segmented genome of eight gene segments, each deposited under a separate accession. Counting accessions therefore overcounts viruses up to eight-fold; we deduplicated to the *isolate* level using the “`/strain`” qualifier.

During validation we observed that some laboratories embed the segment name within the “`/strain`” field itself (e.g., `A/chicken/Bangladesh/WGS-11-2025-PB2`), which causes strain-level deduplication to treat the eight segments of one virus as up to eight distinct isolates. We therefore canonicalized strain identifiers by stripping a trailing segment token (one of the eight segment names or their standard abbreviations) before deduplication. This refinement of the preregistered deduplication rule affected one laboratory's submissions in a single country (Bangladesh) and reduced that country's isolate count from 923 to 814; no other country was affected. The refinement is a documented deviation from the Open Science Framework preregistration (`https://osf.io/dcx3e`).

#### 3. Subtype and clade classification

Each record was classified as H5 or non-H5 from the "/serotype" qualifier, using the organism name and sequence definition as fallback where the serotype qualifier was absent. GenBank serotype encodes subtype (e.g., H5N1) but not pathotype (highly vs low pathogenic); we retained all H5 subtypes and ran a sensitivity analysis excluding the likely low-pathogenic H5N2/H5N3 records. Nineteen isolates carried these subtypes (16 H5N2, 3 H5N3), distributed only in Bangladesh (11) and Myanmar (8); no other country, India included, held any. Excluding them reduced the regional total from 919 to 900 isolates, left the geographic concentration essentially unchanged (Bangladesh 803 of 900, 89.2%, versus 88.6% with all subtypes) with every country's rank preserved, and did not alter India's coverage index (42 isolates; 13.0 per 100 under either definition); the only material change was Myanmar (18 to 10 isolates), whose high outbreak-based ratio the main analysis already attributes to sparse reporting rather than dense sequencing. Hemagglutinin clade is not reliably resolvable from GenBank deposition metadata and was not assigned to the open-access records. For the GISAID subset we summarized the database's curated hemagglutinin clade field.

#### 4. Genome completeness categories

For each isolate we counted the number of distinct gene segments deposited (1–8) and recorded whether the PB2 segment (which carries the mammalian-adaptation markers PB2 E627K and D701N) was present. For display (Figure 3), completeness was binned as: single segment (1), few (2–3), partial (4–7), and whole genome (≥8, i.e., all segments present). The whole-genome secondary coverage metric counted only isolates with all eight segments.

#### 5. Host classification

Host was classified from the free-text "/host" qualifier into six categories: domestic poultry; wild bird; genus-level *Anas* sp. waterfowl of unresolved domestication status; other avian; mammal; and environment. The *Anas* sp. category (predominantly from Bangladesh's domestic-duck surveillance) was retained separately rather than assigned to a wild-versus-domestic class, because genus-level labels do not resolve domestication status. Metadata completeness was computed as the proportion of isolates carrying an annotated host and a day-level collection date.

#### 6. GISAID integration and cross-deduplication

For the restricted-database robustness analysis we exported all influenza A(H5) isolates (any neuraminidase subtype, any host) collected during 2015–2025 for the nine countries from GISAID EpiFlu, deduplicated them by EPI_ISL accession, and cross-deduplicated against the GenBank isolates by canonical strain within country so that a virus deposited to both databases was counted once. Combined coverage was recomputed over the unchanged outbreak denominator. Consistent with the GISAID data-sharing terms, only aggregate per-country counts are reported and isolate-level GISAID records were not redeposited; the corresponding data set is cited as EPI_SET_260705su (`doi:10.55876/gis8.260705su`) with its acknowledgement table.

#### 7. Coverage index and confidence intervals

The coverage index (unique H5 isolates per 100 reported H5 outbreak events) combines two independently enumerated surveillance streams; the numerator is not a subset of the denominator, the index is unbounded, and it is undefined where no outbreaks are reported. Exact Poisson (gamma-method) 95% confidence intervals were computed on the isolate count and propagated to the index with the reported-outbreak denominator held fixed; the intervals therefore reflect the discreteness of the genomic count and, by construction, not the systematic reporting uncertainty in the denominator.

#### 8. WOAH WAHIS denominator cross-check

To triangulate the EMPRES-i denominator against its primary source, we retrieved the complete WOAH WAHIS epidemiological-event record and filtered to H5 avian influenza events in the nine countries for 2015–2025 (`script 12_pull_wahis.py`). WAHIS returned 73 H5 events for India, 14 for Nepal, 8 for Bangladesh, 7 for Bhutan, 4 each for Myanmar, Pakistan, and Afghanistan, and 0 for Maldives and Sri Lanka; the rank order matched EMPRES-i almost exactly (Spearman ρ = 0.99). WAHIS epidemiological events are a coarser counting unit than the EMPRES-i event records, and WAHIS is the notification stream EMPRES-i ingests, so the two are neither comparable in magnitude nor fully independent; the cross-check validates the concordant ranking, not the absolute totals.

#### 9. Sensitivity to the outbreak denominator

The coverage index depends on the reported-outbreak denominator, so we recomputed it (`script 15_denominator_sensitivity.py`) under three denominator definitions with the isolate numerator held fixed: EMPRES-i outbreak *events* (primary), EMPRES-i outbreak *observations*, and WOAH WAHIS epidemiological *events* (Section 8). The seven events whose year was recovered from the report date, where the observation date was missing, are retained in these counts; they are too few (seven of several hundred, none altering India's position as the highest-burden country) to change any country's rank or the coverage conclusions. In this export the EMPRES-i event and observation counts coincide for every country, so the substantive contrast is EMPRES-i versus WAHIS. The country rank order of the index was preserved across denominators (Spearman ρ = 0.99 between the EMPRES-i and WAHIS indices; ρ = 1.00 between EMPRES-i events and observations): Bangladesh remained the extreme high outlier under all three (≥ 4,500 isolates per 100 events), while the two highest-burden countries, India and Nepal, remained far below it (India 13.0 per 100 under either

EMPRES-i denominator and 57.5 per 100 under the smaller WAHIS denominator). Absolute index values rise under the coarser WAHIS denominator, but the mismatch between outbreak burden and genomic coverage is denominator-invariant (Appendix Table A1; Appendix Figure A1, `figS_denominator_sensitivity.png`).

**Appendix Table A1.** Coverage index (unique open-access H5 isolates per 100 reported H5 events) under three outbreak-denominator definitions. Index undefined where no outbreaks were reported.

| Country | Isolates | Index/100 (EMPRES-i events) | Index/100 (EMPRES-i obs.) | Index/100 (WAHIS events) |
|---|---|---|---|---|
| India | 42 | 13.0 | 13.0 | 57.5 |
| Nepal | 2 | 2.6 | 2.6 | 14.3 |
| Bangladesh | 814 | 4522.2 | 4522.2 | 10175.0 |
| Bhutan | 1 | 11.1 | 11.1 | 14.3 |
| Myanmar | 18 | 225.0 | 225.0 | 450.0 |
| Pakistan | 17 | 212.5 | 212.5 | 425.0 |
| Afghanistan | 0 | 0.0 | 0.0 | 0.0 |
| Maldives | 16 | — | — | — |
| Sri Lanka | 9 | — | — | — |

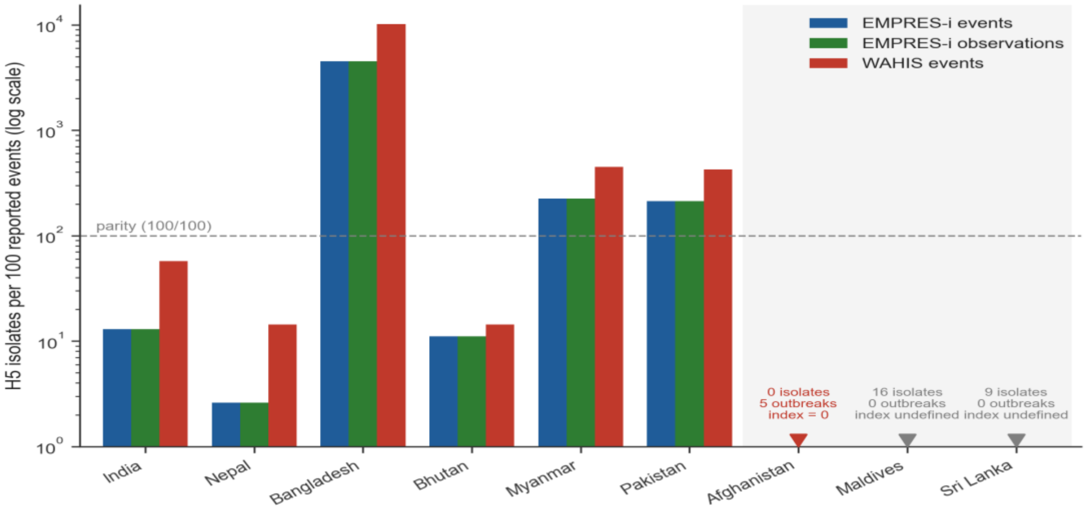


**Appendix Figure A1.** Coverage index (H5 isolates per 100 reported events, log scale) under the three outbreak-denominator definitions, shown for all nine countries. The country rank order is preserved and Bangladesh remains the extreme high outlier under all three. Three countries have no bar because the coverage index cannot be expressed as a value for them and are instead shown as annotated markers at the axis floor: Afghanistan reported outbreaks but deposited no open-access isolates (index = 0), whereas Maldives (16 isolates) and Sri Lanka (9 isolates) have open-access isolates but no reported outbreaks in the study window, leaving the index undefined.

### 10. Temporal robustness of the coverage gap

To test whether the concentration and coverage gap reflect a single outbreak year, for example the post-2020 clade 2.3.4.4b panzootic, we split the study period into 2015–2019 and 2020–2025 and recomputed the numerator, denominator, and index per country per period (`script 14_temporal_trend.py`; Appendix Table A2; Appendix Figure A2, `figS_temporal_trend.png`). The pattern was stable across both periods. Bangladesh accounted for 95.0% of open-access isolates in 2015–2019 and 84.7% in 2020–2025. India's coverage index remained an order of magnitude below Bangladesh's in both periods (5.2 versus 2,500 per 100 in 2015–2019; 15.5 versus 9,780 per 100 in 2020–2025); notably, India's reported outbreak burden more than tripled between periods (77 to 245 events) while its isolate count stayed low (4 to 38 isolates), so the gap is a persistent structural feature of regional surveillance rather than a one-year artifact.

**Appendix Table A2.** Unique open-access H5 isolates, reported H5 outbreak events, and the coverage index by country for the two study sub-periods. Index undefined where no outbreaks were reported.

| Country | Isolates 2015–19 | Isolates 2020–25 | Events 2015–19 | Events 2020–25 | Index/100 2015–19 | Index/100 2020–25 |
|---|---|---|---|---|---|---|
| India | 4 | 38 | 77 | 245 | 5.2 | 15.5 |
| Nepal | 0 | 2 | 24 | 54 | 0.0 | 3.7 |
| Bangladesh | 325 | 489 | 13 | 5 | 2500.0 | 9780.0 |
| Bhutan | 1 | 0 | 5 | 4 | 20.0 | 0.0 |
| Myanmar | 4 | 14 | 8 | 0 | 50.0 | — |
| Pakistan | 8 | 9 | 3 | 5 | 266.7 | 180.0 |
| Afghanistan | 0 | 0 | 4 | 1 | 0.0 | 0.0 |
| Maldives | 0 | 16 | 0 | 0 | — | — |
| Sri Lanka | 0 | 9 | 0 | 0 | — | — |

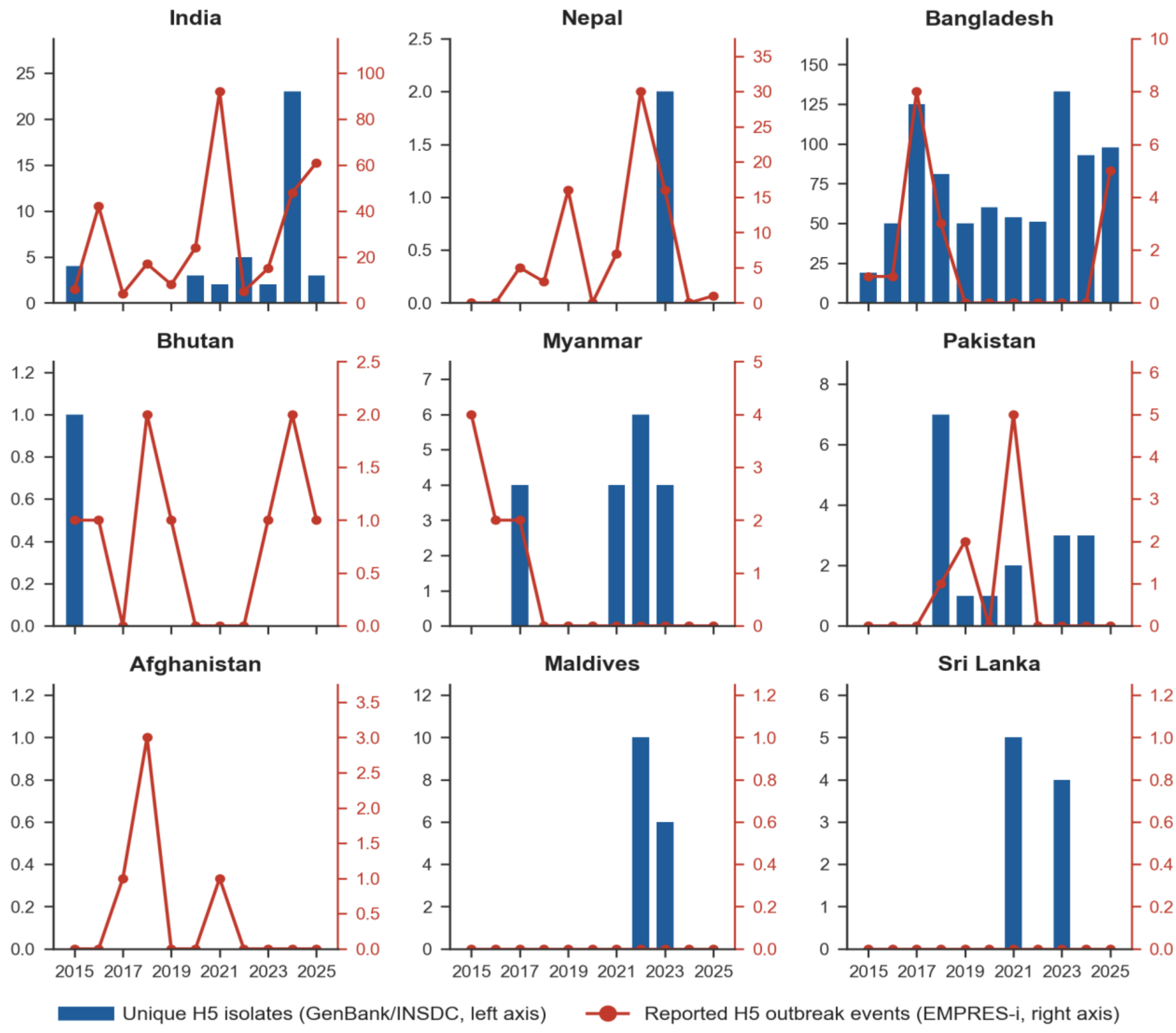


**Appendix Figure A2.** Unique open-access H5 isolates (bars, left axis) and reported H5 outbreak events (line, right axis) by year, 2015–2025, for each country. Axes are scaled independently per panel; the separation between outbreak burden and genomic output persists across both sub-periods.

### 11. Sequencing deficit relative to reported burden

To express the gap in absolute terms we estimated, for each country, the number of genomic isolates expected at a benchmark sequencing rate given its reported outbreak burden, and the shortfall relative to the observed count (`script 16_sequencing_deficit.py`; Appendix Table A3; Appendix Figure A3, `figS_sequencing_deficit.png`). Three transparent benchmarks are reported side by side: parity (one isolate per reported outbreak event, a minimal aspiration), one-in-ten (0.1 per event, the lower guide line in Figure 2), and Bangladesh's observed within-region rate (45.2 isolates per event). At parity, India shows a shortfall of 280 isolates (42 observed versus 322 expected; an 87% deficit), and the region excluding Bangladesh a shortfall of 350 isolates (80 observed versus 430 expected). We note that against the weakest benchmark (one-in-ten) India's observed rate (0.13 per event) is already met, so the deficit is stated against parity rather than the least demanding threshold; benchmarking against Bangladesh's rate is reported only to illustrate the within-region ceiling and is not proposed as a normative target.

**Appendix Table A3.** Observed isolates versus the count expected at parity (one isolate per reported outbreak event) and the resulting deficit, with the deficit against Bangladesh's within-region rate (45.2 isolates/event) shown for reference. Deficit undefined where no outbreaks were reported.

| Country | Observed | Events | Expected at parity | Deficit at parity | Deficit % at parity | Deficit at Bangladesh rate |
|---|---|---|---|---|---|---|
| India | 42 | 322 | 322 | 280 | 87.0 | 14520 |
| Nepal | 2 | 78 | 78 | 76 | 97.4 | 3525 |
| Bangladesh | 814 | 18 | 18 | 0 | 0.0 | 0 |
| Bhutan | 1 | 9 | 9 | 8 | 88.9 | 406 |
| Myanmar | 18 | 8 | 8 | 0 | 0.0 | 344 |
| Pakistan | 17 | 8 | 8 | 0 | 0.0 | 345 |
| Afghanistan | 0 | 5 | 5 | 5 | 100.0 | 226 |
| Maldives | 16 | 0 | — | — | — | — |
| Sri Lanka | 9 | 0 | — | — | — | — |
| **Region excl. Bangladesh** | **80** | **430** | **430** | **350** | **81.4** | **19366** |

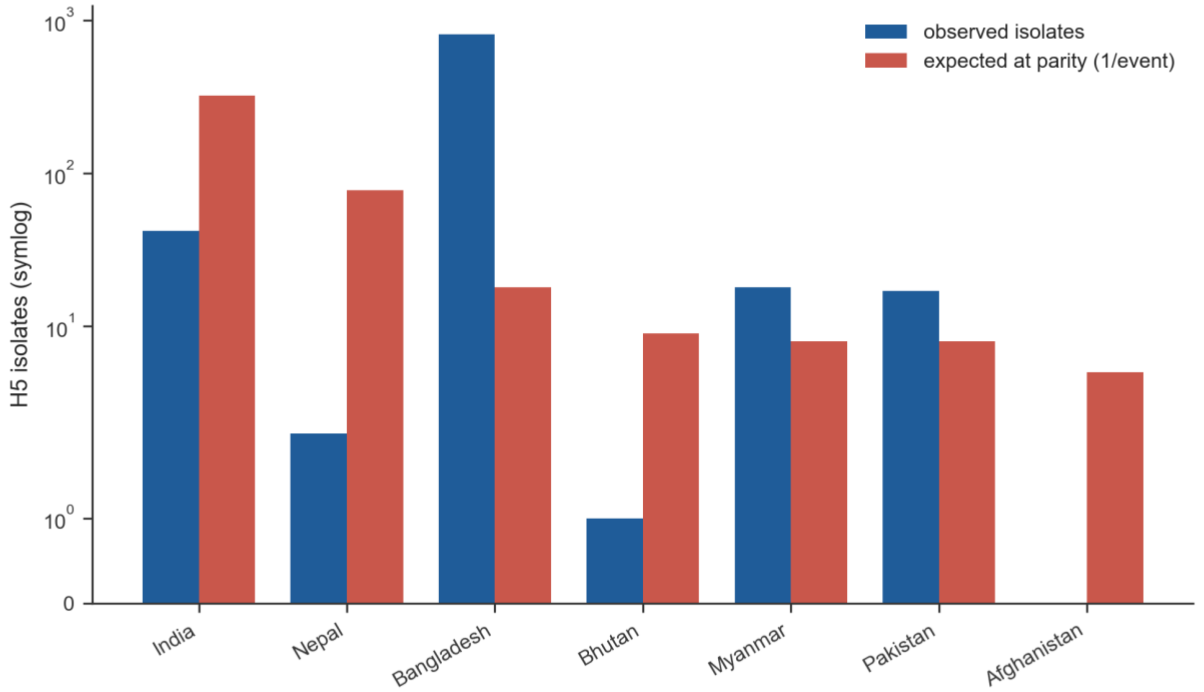


**Appendix Figure A3.** Observed isolates versus the number expected at parity (one isolate per reported outbreak event; symmetric-log scale). India and Nepal fall far below parity, whereas Myanmar and Pakistan exceed it; countries with no reported outbreaks are omitted.

#### 12. Deposition timeliness (collection to public availability)

Surveillance value depends not only on how many genomes exist but on how quickly they become public: a genome released long after collection cannot inform the outbreak response it documents. For every isolate in the numerator we retrieved the GenBank first-public-release date (GBSeq_create-date) by a targeted re-fetch of its segment accessions (`script 13_deposition_lag.py`; the fetched dates are cached with the archive for offline reproduction). An isolate's availability date was taken as the earliest release date among its segments, and the lag computed as release date minus collection date. The metric was restricted to the 880 of 919 isolates whose collection date was month-or-finer (39 year-only dates were excluded as too coarse; no isolate had a negative lag). The create-date is the date the record first became publicly retrievable from GenBank, not the earlier submission date or the date of any associated publication, so the lag is a conservative measure of how long each sequence was unavailable for reuse.

Deposition was slow across the region: the median collection-to-availability lag was 392 days (interquartile range 238–762 days), i.e. roughly 13 months (IQR ~8–25 months). The lag was long even in the two countries that sequence at scale, Bangladesh median 377 days (~12.4 months; n = 814) and India median 494 days (~16.2 months; n = 42), and longer still, though on small counts, for Pakistan (811 days; n = 17), Myanmar (545 days; n = 4) and Nepal (1,116 days; n = 2); Afghanistan, Maldives and Sri Lanka contributed no dated isolates. Because the great majority of open-access H5 genomes from the region reach the public record more than a year after the sampled event, the open-access record functions as a retrospective archive rather than a real-time surveillance signal, a timeliness gap that compounds the volume and completeness gaps documented above (Appendix Table A4; Appendix Figure A4, `figS_deposition_lag.png`). The release dates were strongly clustered, the signature of retrospective batch deposition rather than continuous submission: the 880 dated isolates became public on only 98 distinct dates, and 74% were released in bulk groups of ten or more sharing an identical release date. The largest single batch comprised 91 isolates released together on one day, a median 5.2 years after collection, so the long right tail of the distribution reflects sequences deposited in publication-linked bulk drops rather than a uniform per-isolate delay.

**Appendix Table A4.** Collection-to-public-availability lag by country, restricted to the 880 of 919 isolates with a month-or-finer collection date. n = isolates contributing; medians on very small n (Bhutan, Nepal, Myanmar) are shown for completeness only.

| Country | n | Median (days) | IQR (days) | Median (months) |
|---|---:|---:|---:|---:|
| India | 42 | 494 | 166–594 | 16.2 |
| Nepal | 2 | 1116 | 1106–1126 | 36.7 |
| Bangladesh | 814 | 377 | 238–752 | 12.4 |
| Bhutan | 1 | 161 | 161–161 | 5.3 |
| Myanmar | 4 | 545 | 545–545 | 17.9 |
| Pakistan | 17 | 811 | 40–1027 | 26.6 |
| Afghanistan | 0 | — | — | — |
| Maldives | 0 | — | — | — |
| Sri Lanka | 0 | — | — | — |
| **All** | **880** | **392** | **238–762** | **12.9** |

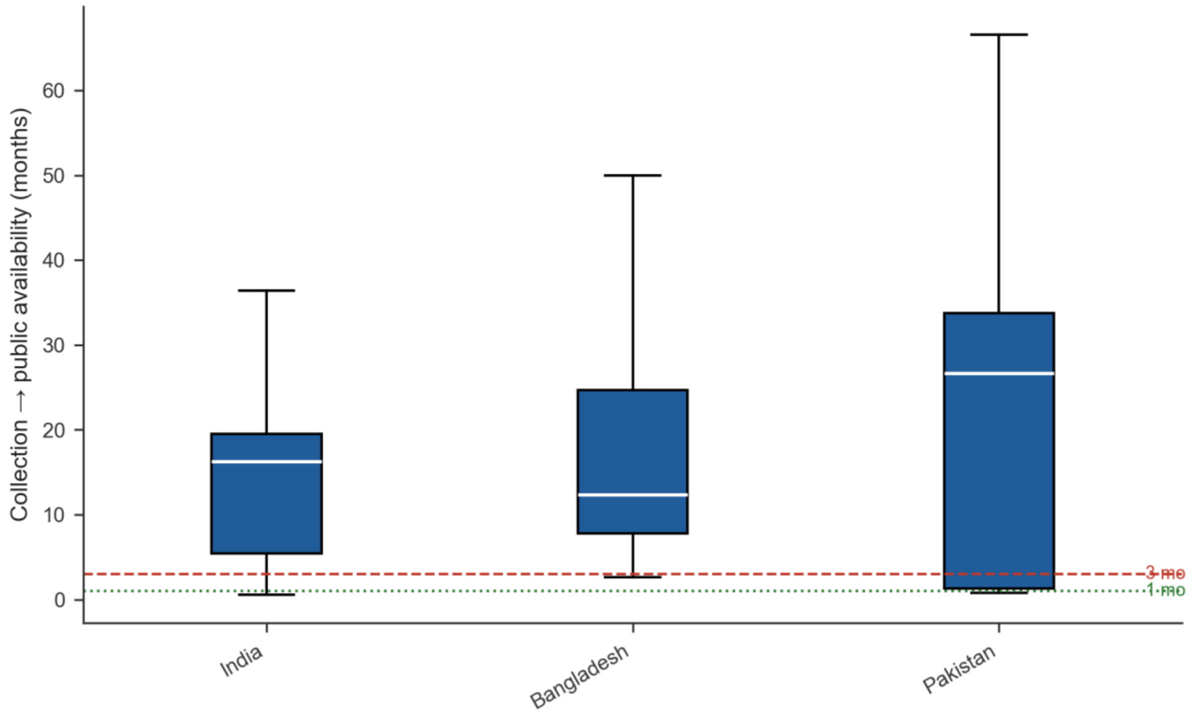


**Appendix Figure A4.** Distribution of the collection-to-public-availability lag (months) for the countries with a sufficient number of dated isolates (n ≥ 10: India, Bangladesh, Pakistan; smaller-n countries are reported in Table A4 only). Dotted and dashed lines mark 1-month and 3-month reference thresholds; all three national medians exceed one year.

### 13. Formal quantification of geographic concentration

The concentration of open-access genomic output is the study's principal denominator-free result, so we quantified geographic inequality in isolate counts using the Gini coefficient, alongside the top-country and top-two shares (script 17_concentration_index.py; Appendix Table A5; Appendix Figure A5). The Gini coefficient was calculated across all nine countries, including the country with no deposited isolates, and does not depend on the reported-outbreak denominator or its reporting bias. Open-access output was extremely unequal (Gini coefficient = 0.83), with the single top country holding 88.6% and the top two 93.1% of the regional record. The result was unchanged on the combined GenBank ∪ GISAID numerator (Gini coefficient = 0.83), confirming that it is not an artifact of restricting the analysis to open-access data.

**Appendix Table A5.** Geographic inequality in H5 isolate deposition across the nine countries, calculated from isolate counts alone (independent of the outbreak denominator).

| Metric | Open-access (GenBank) | Combined (GenBank ∪ GISAID) |
|---|---|---|
| Contributing countries (of 9) | 8 | 8 |
| Total isolates | 919 | 1,297 |
| Top-country share | 88.6% | 89.3% |
| Top-two share | 93.1% | 92.8% |
| Gini coefficient | 0.827 | 0.826 |

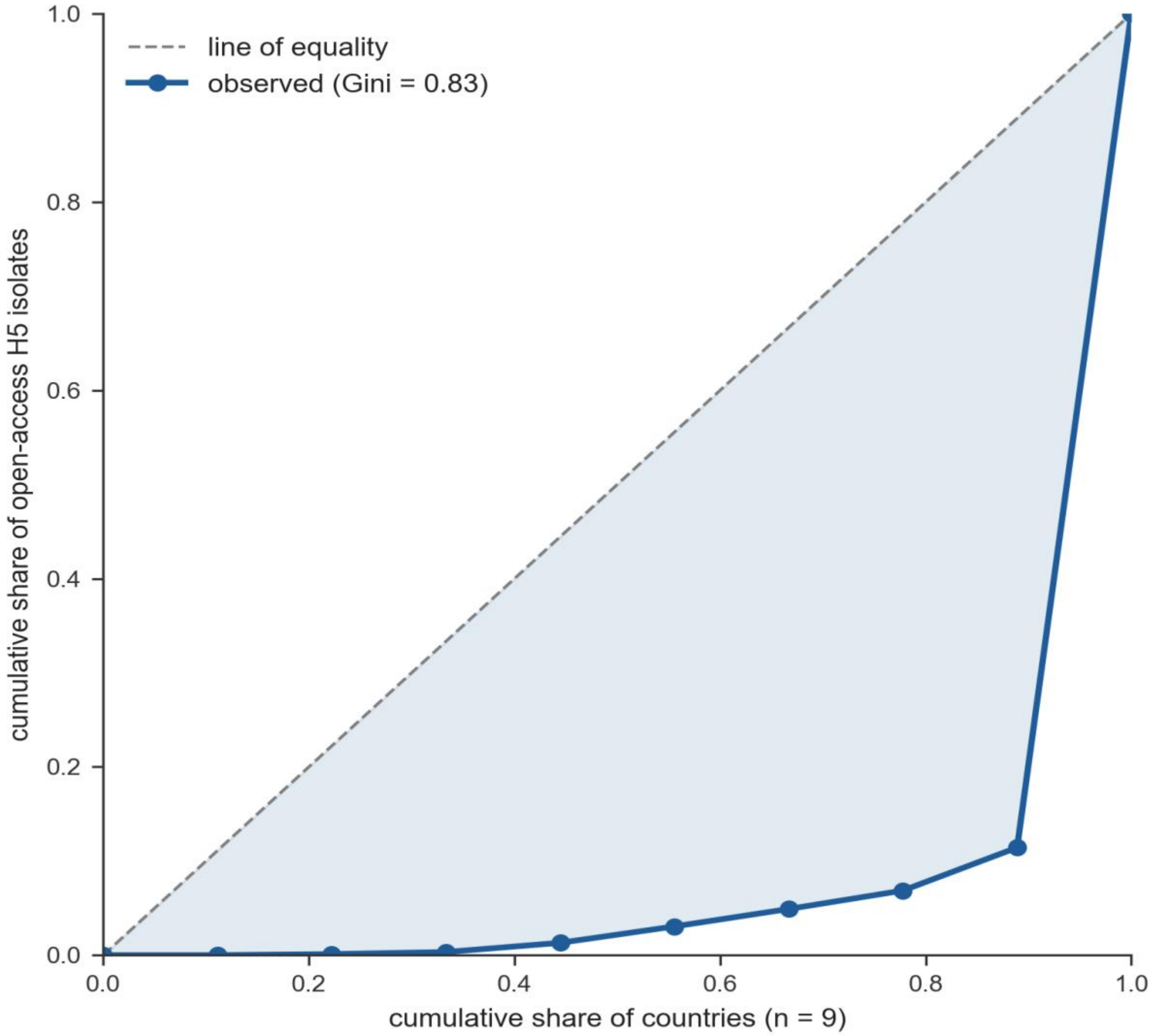


**Appendix Figure A5.** Lorenz curve of open-access H5 genomic output across the nine countries. The dashed diagonal is the line of perfect equality (each country contributing an equal share); the observed curve hugs the axis and rises almost vertically at the final country, the geometric signature of extreme concentration (Gini coefficient = 0.83).

#### 14. Coverage under reporting-bias-free denominators

Every coverage ratio built on reported outbreaks inherits the objection that reported outbreaks track reporting *effort* rather than true burden. To test the pattern against denominators enumerated entirely independently of animal-health reporting, we re-expressed genomic output per unit of poultry reservoir and per capita (`script 18_independent_denominator.py`; Appendix Table A6; Appendix Figure A6). Poultry stocks (chickens, ducks, turkeys, and geese combined) came from the FAO live-animal "Poultry – Stocks (animals)" series (latest year, 2024; retrieved via Our World in Data) and human population from the World Bank (SP.POP.TOTL, 2023); both extracts are archived with the analysis code. Because poultry stocks are a single-year snapshot (2024) whereas the numerator accumulates over 2015–2025, the reservoir denominator is a scale proxy rather than a cumulative-exposure measure; national flock sizes are stable enough at this order of magnitude that the ranking of the large-reservoir countries (Bangladesh, India, Pakistan) is unaffected. Maldives has no FAO poultry-stocks record (negligible island poultry), so its poultry-normalized coverage is undefined, as its outbreak index already is.

The concentration and India's deficit persisted under both independent denominators. Bangladesh sequenced far more per unit reservoir than any other country (2,055 isolates per billion poultry and 47 per 10 million people), confirming that its dominance is a genuine difference in sequencing intensity and not merely a low-reporting artifact of the outbreak denominator. India (47 isolates per billion poultry) and, most strikingly, Pakistan (8 per billion poultry) sat near the floor despite holding the region's two largest flocks (887 million and 2.07 billion birds). Pakistan is the clearest case the reservoir denominator exposes: its outbreak index (212 per 100) makes it appear well characterized, but that figure reflects only eight reported outbreaks, and against its enormous reservoir Pakistan is the most under-sampled country in the region. The reservoir view therefore adds information rather than restating the outbreak index (Spearman $\rho$ between the two = 0.64 for poultry and 0.71 for population). Per-unit rates are unstable for the smallest reservoirs and populations (Bhutan, Sri Lanka, and Maldives contribute 1–16 isolates over very small denominators) and should be read only for the large-reservoir countries, where the pattern is robust.

**Appendix Table A6.** Genomic output per unit poultry reservoir and per capita, with the reported- outbreak coverage index shown alongside for comparison. Poultry stocks: FAO, 2024. Population: World Bank, 2023. Per-unit rates on very small denominators (Bhutan, Maldives, Sri Lanka) are shown for completeness only.

| Country | Isolates | Poultry stocks (2024) | Isolates / billion poultry | Population (2023) | Isolates / 10M people | Outbreak index /100 |
|---|---|---|---|---|---|---|
| India | 42 | 886,518,000 | 47.4 | 1,438,069,596 | 0.29 | 13.0 |
| Nepal | 2 | 57,713,000 | 34.7 | 29,694,614 | 0.67 | 2.6 |
| Bangladesh | 814 | 396,038,000 | 2055.4 | 171,466,990 | 47.47 | 4522.2 |
| Bhutan | 1 | 924,000 | 1082.3 | 786,385 | 12.72 | 11.1 |
| Myanmar | 18 | 153,095,000 | 117.6 | 54,133,798 | 3.33 | 225.0 |
| Pakistan | 17 | 2,067,907,000 | 8.2 | 247,504,495 | 0.69 | 212.5 |
| Afghanistan | 0 | 13,480,000 | 0.0 | 41,454,761 | 0.00 | 0.0 |
| Maldives | 16 | — | — | 525,994 | 304.19 | — |
| Sri Lanka | 9 | 18,732,000 | 480.5 | 22,037,000 | 4.08 | — |

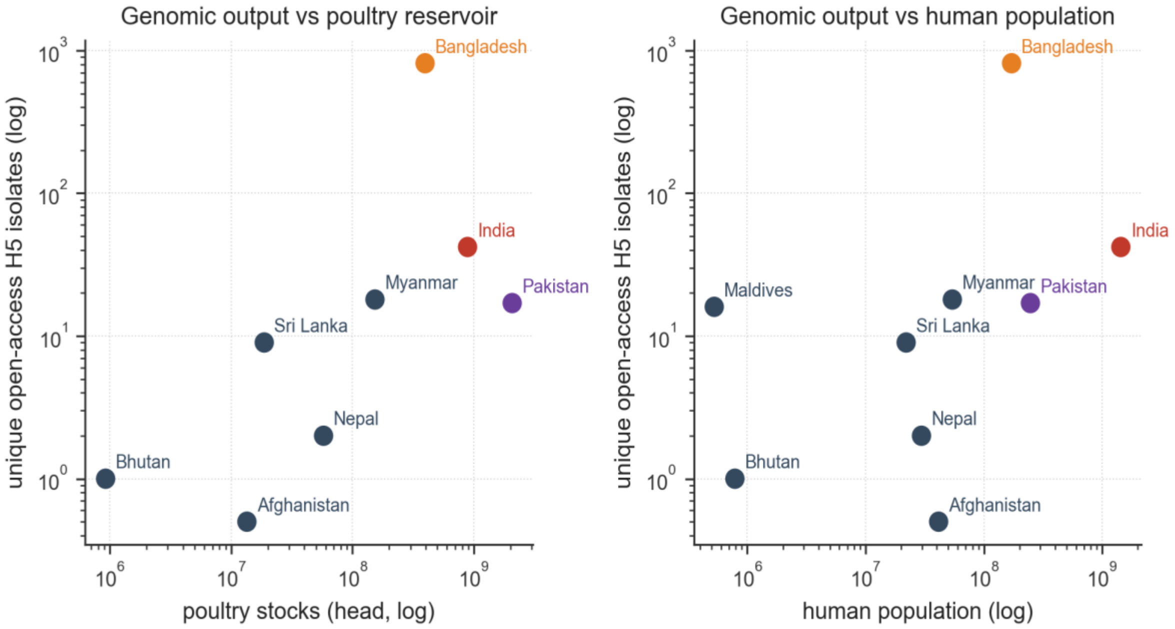


**Appendix Figure A6.** Open-access H5 isolates versus poultry reservoir size (left) and human population (right), each country one point on logarithmic axes. Bangladesh (orange) lies far above the regional scatter under both denominators, whereas India (red) and Pakistan (purple), the two largest poultry reservoirs, lie far below it, so the coverage gap is not an artifact of the outbreak-reporting denominator. Afghanistan (0 isolates) is plotted at a display floor of 0.5; Maldives is omitted from the poultry panel (no FAO poultry record).

#### 15. Software environment

All data retrieval, processing, statistical analysis, and figure generation were performed in `Python 3.13.7`. The analysis depended on the following packages, pinned to the exact versions used to produce every reported statistic and figure: `Biopython 1.87`, `pandas 3.0.2, NumPy 2.4.4, SciPy 1.17.1, Matplotlib 3.10.9, Pillow 12.2.0, openpyxl 3.1.5, xlrd 2.0.2`, and `certifi 2026.4.22`. These pinned versions are archived with the analysis code at Zenodo to support byte-identical reproduction of the reported results.
